# Modeling the role of clusters and diffusion in the evolution of COVID-19 infections during lock-down

**DOI:** 10.1101/2020.07.22.20159830

**Authors:** W. J. T. Bos, J.-P. Bertoglio, L. Gostiaux

## Abstract

Epidemics such as the spreading of the SARS-CoV-2 virus are highly non linear, and therefore difficult to predict. In the present pandemic as time evolves, it appears more and more clearly that a clustered dynamics is a key element of description. This means that the disease rapidly evolves within spatially localized networks, that diffuse and eventually create new clusters. We improve upon the simplest possible compartmental model, the SIR model, by adding an additional compartment associated with the clustered individuals. This sophistication is compatible with more advanced compartmental models and allows, at the lowest level of complexity, to leverage the well-mixedness assumption. The so-obtained SBIR model takes into account the effect of inhomogeneity on epidemic spreading, and compares satisfactorily with results on the pandemic propagation in a number of European countries, during and immediately after lock-down. Especially, the decay exponent of the number of new cases after the first peak of the epidemic is captured without the need to vary the coefficients of the model with time. We show that this decay exponent is directly determined by the diffusion of the ensemble of clustered individuals and can be related to a global reproduction number, that overrides the classical, local reproduction number.

---

Perhaps the most widely used model in epidemiology is the SIR model, dividing the population into three compartments (Susceptible-Infected-Removed). A well known flaw of this approach is the underlying assumption that all susceptible individuals are in contact with the infected individuals, thereby ignoring the spatial character of the epidemic. We introduce a two-scale model, where susceptible individuals are either in the vicinity, or spatially distant from the infected. The interaction between these two compartments is inspired by a diffusion process. Different time scales govern the spatial spreading of the epidemic and the local increase of the number of infections. Thereby, the epidemic can reach a peak corresponding to local collective immunity well before global immunity is attained. It is shown, comparing to data of the COVID-19 epidemic for various European countries, that the present model, with constant model-coefficients, reproduces quite precisely the initial growth phase of the epidemic and the decrease of the number of infections during lock-down.

## I. LOCK-DOWN AND MODELING OF THE COVID-19 EPIDEMIC

Since the beginning of the COVID-19 epidemic in december 2019, many countries have chosen for severe restrictions on mobility and transport. It has become a controversial question in public debate whether these decisions were necessary and inevitable, and whether the lock-down was maintained for an unnecessary long period or, on the contrary was relaxed too soon in an unsafe way. In order to evaluate retrospectively such decisions, and to anticipate similar measures in the future, an in-depth understanding of epidemic propagation is needed. In the present investigation we focus on the seemingly universal way in which the SARS-CoV-2 disease is controlled by lock-down.

Indeed, in a number of countries in which lock-down was applied, the number of new infections decays exponentially after the peak of the epidemic with a very similar time-scale (see Fig. 3). In the present investigation we shed light on the role of the finite size of spatially localized clusters on this phenomenon, by improving upon a simple paradigm model, the classical SIR model, and by assessing the resulting SBIR model against realistic data.

The SIR model in its simplest form contains three compartments, corresponding to susceptible, infected and recovered or removed individuals, and the dynamics are governed by two model parameters *γ* (recovery rate) and *β* (infection rate). A wide variety of SIR modifications have seen the light since its original formulation^1^. Many of them propose additional compartments (see for instance Ref. 2 for an analysis of a great number of compartmental models), nonlinear incidence rates^3^, and non-constant model coefficients^4,5^. An overview of these models is beyond the scope of the present work and we refer to the review articles on the subject^6,7^.

In a large number of these epidemiologic studies using compartmental models, the dynamics are extremely sensitive on the temporal variations of the model parameters, and saturation of the epidemic at a realistic level is only obtained if the main parameters, such as the infection rate (or reproduction number *ℛ*_0_ = *β/γ*) are changed during the analysis. Sophisticated approaches have been developed to estimate the temporal evolution of *β* by assuming for instance a prescribed shape of the time-evolution of the reproduction number^8^ or by adjusting the value of *β* instantaneaously to the observations^5^. We show that even using constant coefficients, saturation of the epidemic is obtained when the local nature of interactions within clusters is taken into account.

Indeed, perhaps the largest known flaw of the SIR model is the assumption of well mixedness. By assimilating all susceptible individuals into one single compartment, it is tacitly assumed that all individuals are in contact with all others, and the spatial structure of the epidemic is thereby ignored. An approach to overcome this weakness is to use either networks, self-propelled interacting agents, nested or coupled local SIR-type models^9^ or diffusion equations^10–12^. Such modifications change the level of complexity of the model considerably requiring either the solution of a two-dimensional reaction-diffusion problem, tracking a large number of propelled particles, or the solution of integro differential equations. We refer to Ref. 13 for a discussion of the differences between agent-based and compartmental models.

The model we introduce in the present work is much simpler and uses a two-scale approach: either individuals are far enough away from the individuals not to be in contact, or they pertain to a cluster of individuals in contact with the infected. This approach allows to take into account spatial structuring at the lowest level of complexity, i.e., by adding one single compartment. We sub-divide the group of susceptibles into *B*, those in a so-called “blob”, who are or have been in contact with the infected, and *S*, those who are (spatially) distant from the infected. We will show, by comparison with data for a variety of European countries, that this model reproduces the exponential decay of the number of newly infected after its peak-value, with an exponent determined by the spatial diffusion coefficient of the blob.

An important simplification of reality which we will assume throughout this investigation is the constancy of the model-coefficients. All countries introduced different measures which change the effective transmission rate by physical effects such as quarantine-measures^14^, or by changing the awareness of the individuals^15^. The present investigation aims primarily at the assessment of a novel approach to take into account inhomogeneity. The lock-down period for a number of similar occidental countries seems the correct starting point, since despite differences in detailed measures, the parameters governing the dynamics of the epidemic are expected to become constant during lock-down. The universal features captured by the model can therefore be disentangled from other effects, and the results can be compared to the data within the limits of the quality of the measurements.

In the following we will first present the SBIR model and its difference with the classical SIR approach (section II). We will analytically determine the exponential decay time-scale for the SIR model with and without cross-immunity, as well as for the SBIR model. We show, by comparison with data from the Johns Hopkins University data-base^16^, that the SBIR model allows to simultaneously fit the data in the increasing and decreasing phase of the epidemic without the need of adjusting the model-parameters during the epidemic (section III). We further estimate the reproduction number and show that it reaches a stable minimal value during lock-down. The appendices contain the mathematical procedure to analytically estimate the decay exponents of the different models and the initial and final reproduction numbers.

## II. A CLUSTER-BASED COMPARTMENTAL MODEL

Spatially resolved epidemiological models are elegant and physically better justified than the SIR model but have the disadvantage that they are harder to analyze analytically, which makes insights on the effect of the different parameters more complicated. Indeed the complexity increases in general the descriptive capacities of a model, but decreases the insights which can be obtained.

We propose here a new model, that does only introduce one new compartment, representing the individuals who are or have been spatially close to the infected people. Thereby our model remains based on global parameters and does not introduce the need to solve a two-dimensional diffusion problem. This simplicity allows the analytical derivation of the long-time behavior and thereby elucidates the role of local cluster-saturation on global decay-rates.

We will in this section first recall the SIR model, give an analytical estimate of the decay-exponent in the final phase of the epidemic, and show the influence of cross-immunity in a simple CSIR model. After that we introduce the SBIR model and discuss its physical properties.

### A. The decay exponent in SIR and cross-immunity

The simplest model for macroscopic modeling of an epidemy is the so-called SIR model^1,17^. The model describes a population divided in three categories, Susceptible *S*, Infected *I* and Removed or Recovered *R* (Fig. 1, top). The total population *N* = *S* + *I* + *R* is supposed constant on the timescale of the epidemic. The evolution of, and the interaction between the three compartments is given by a set of three coupled ordinary differential equations,

**FIG. 1.**
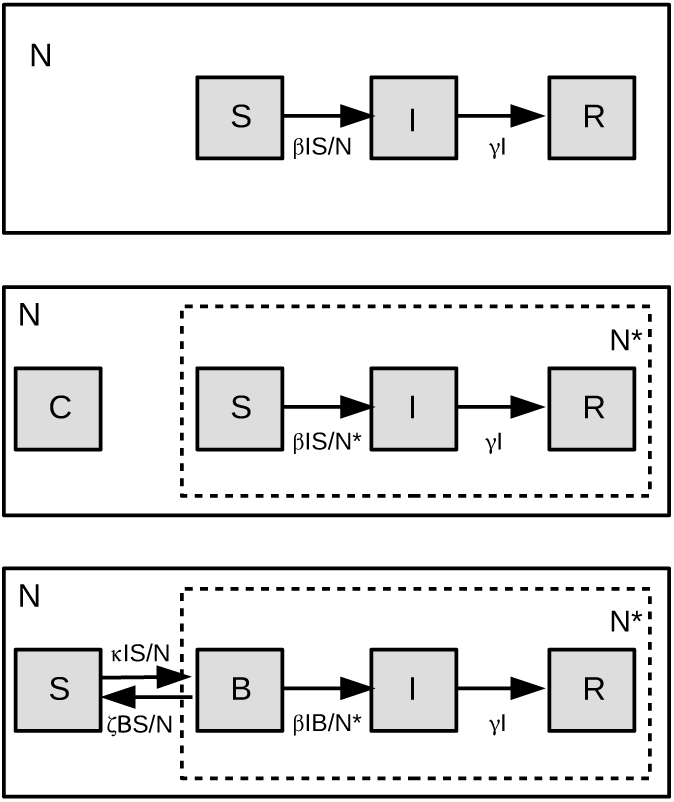
Diagrammatic representation of three compartmental models, SIR, CSIR and SBIR. In SIR and CSIR three compartments interact, with the only difference in CSIR that those three compartments do not contain the entire population, since part of the population is supposed to be protected by cross-immunity. In the SBIR model, the susceptible part of the population is subdivided into individuals *B* inside the blob, and *S*, outside the blob, and the four compartments interact.

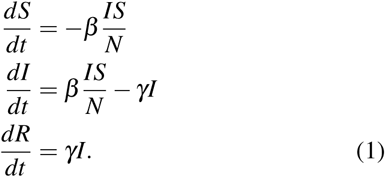

In the following it is convenient to normalize these equations by *N*, so that *R, S, I* all take values between one and zero and *S* + *R* + *I* = *N* = 1. Initially *R*(0) = 0, *I*(0) = *I*_0_ *«* 1 and *S*(0) = 1 − *I*(0) ≈ 1. The dynamics of the SIR model are characterized by an exponential growth with exponent *λ* ^+^ = *β* − *γ*, upto a time *τ* where saturation of the infected is obtained, by collective, or herd immunity, obtained when *S < γ/β*. Note that in the present epidemic of SARS-CoV-2, we remain very far below such collective immunity at the time of the first peak. Long enough after this *t* = *τ*, exponential decay is observed until all infected have disappeared. The behavior is sketched in Fig. 2. We show in appendix A a simple and straightforward way to determine the approximate exponential growth and decay rate of the epidemic. This analysis, assuming continuity of two exponentials (as in Fig. 2), yields the decay-exponent

**FIG. 2.**
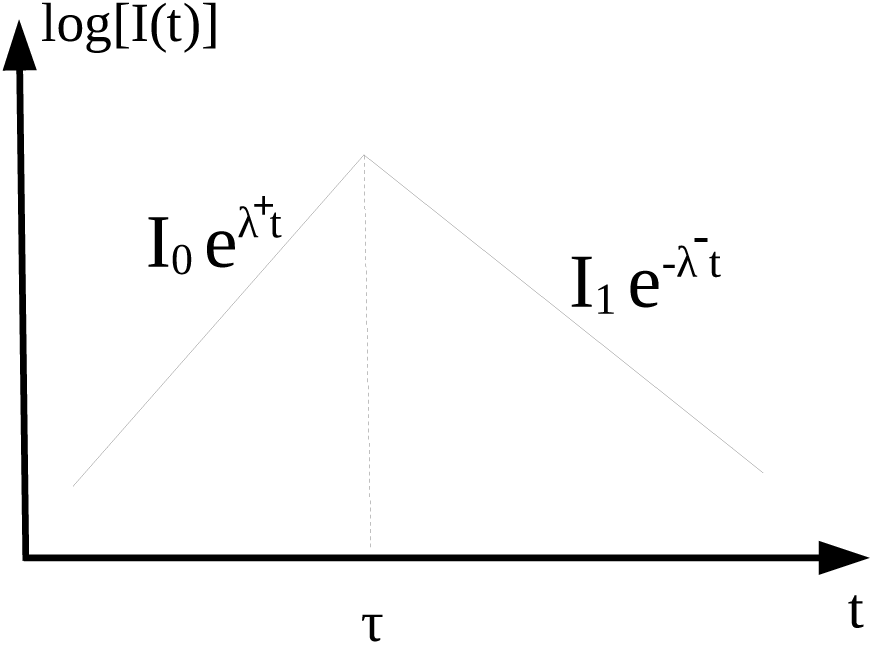
Schematic representation of the evolution of the number of infected individuals in SIR and SBIR under the assumption of constant model-parameters. Time *τ* represents the peak-value of the number of infected.

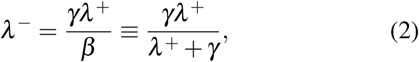

which gives a direct relation between the exponential during the initial, growing phase of the epidemic, and the decaying phase. We note that an exact analytical solution of the system exists^18^, but the extension of the method to more complicated systems might not be straightforward. The present approach yields less accurate but readily interpretable results, allowing direct insights in the effect of the parameters on the evolution of the epidemic.

Furthermore, the peak-value of the number of infections is given by

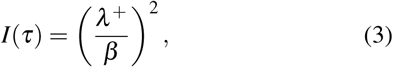

and at this time *τ*, the rate of infection *P* = *β SI* is close to its maximum and is given by (see appendix A)

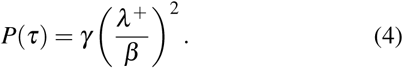

An important question is whether cross-immunity could explain the saturation of an epidemic such as the ongoing one at the observed levels. We consider thereto a population where a part of the individuals has attained cross-immunity - the precise source of it, past contact with similar viruses or any other hidden mechanism being beyond the scope of this study. This part of the individuals is called *C* (Fig. 1, center). To obtain the CSIR equations, the SIR-equations are now only changed on the level of the global community size, where we replace

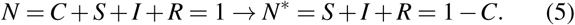

The growth-exponent *λ*^+^ is not changed by this modification for given *γ, β*, but the peak-values of *I* and *P* are multiplied by a factor (1 −*C*) as explained in appendix B. For instance

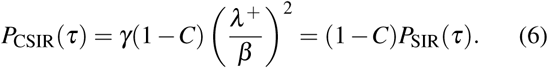

In the long-time however, the decay-exponent *λ*^−^ is the same for the SIR and the CSIR model.

### B. SBIR: SIR in a blob

We consider a cluster-based compartmental model, where only a sub-ensemble of individuals *N*^∗^ *< N* will interact with infected individuals. Via social interaction, a set of clusters diffusing around infected individuals will emerge, this diffusion process being distinct from virus transmission, and having its own dynamics. We define the number of persons belonging to one of these diffusing clusters as *N*^∗^ = *B* + *I* + *R* ≡ 1 − *S*, where *I* and *R* have the same interpretation as before, i.e., infected and recovered/removed, respectively, and *B* is referred to as the blob, namely the people who joined a diffusing cluster through social interaction and who are not yet infected or recovered. These individuals can be seen as local susceptibles, and in the following the *B, I* and *R* compartments are considered globally, independently of the number of clusters they correspond to. The persons in *S* are susceptible individuals, that need first to join the blob before being possibly infected. The 4 compartments, as illustrated in Fig. 1, are now related by the following system of ordinary differential equations,

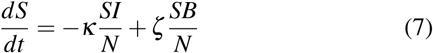

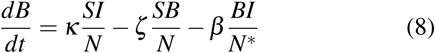

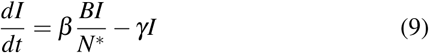

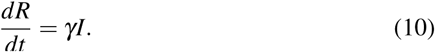

If the growth exponent *λ* ^+^ = *β*− *γ* has the same expression as in the SIR model, the final decay exponent, if *β > κ*, is given by (see appendix C),

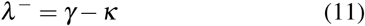

which is different from the expressions for SIR and CSIR, and now depends on the spatial diffusion-coefficient of the blob related to *κ*.

In the limit of very fast diffusion, *B* will approach *S*, and the SIR-dynamics is observed subsequently. Furthermore, mathematical robustness of the formulation is ensured, since none of the quantities *S, B, I, R* can become negative.

### C. The physics behind SBIR

Before integrating the SBIR model, it is insightful to show what the physical picture behind the model is. This will help to interpret the results and to guide the choice of the parameters. The parameters of the model are *γ, β, κ* and *ζ*. Whereas *γ* is an intrinsic parameter characterizing the disease, *β, κ* and *ζ* depend on physical, biological and psychological influences. The *β* parameter, representing the infection rate is already present in the classical SIR model. It will be obtained here from an estimation of *λ* ^+^ in the early stage of the epidemics, which is known to be barely affected by the spatial structure of the population^19^.

In order to understand the dynamics of the blob, let us consider the coupling between the *S* and *B* compartments, and first neglect the term proportional to *ζ*. We have then

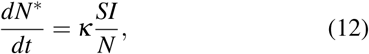

If we assume *S/N ≈* 1 (as expected in early stages) and that *I* varies slowly, we have

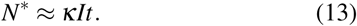

When a constant population density *σ*_*P*_ is introduced, the surface of the blob *N*^∗^*/σ*_*P*_ corresponds to the increase of a surface explored by a random walk. Indeed, the separation of a random walker from its initial position is

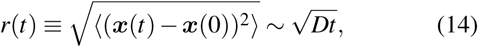

where *D* is the diffusion coefficient, and the surface explored is proportional to the square of this length. The interaction between the two compartments is therefore governed by a Brownian-like diffusion process, with a diffusion coefficient *D* = *κI/σ*_*P*_. In most epidemics, *I* rapidly grows so that the time-varying effective diffusion coefficient increases as well, and super-diffusive growth of the blob is observed.

We can ask how this diffusion is related to the movement of individuals. In the kinetic theory of gases, the macroscopic diffusion coefficient is related to microscopic motion by

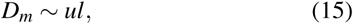

where in this expression *u* is the velocity magnitude of the gas-molecules and *l* the mean-free-path. In our description, where the blob diffuses with a typical timescale *κ*^−1^, the effective mean free path of individuals in contact with infected is proportional to the number of infected *I*, assuming that their velocity, which is related to the displacement of individuals, is constant (and reduced) during lock-down. Even though this is obviously a very rough approximation, we could use this idea to estimate the change of diffusion after lock-down using smartphone mobility data (as for instance^20,21^). In the present investigation we will use constant model-coefficients and show that this approximation, during confinement, gives good agreement with data. The influence of non-constant coefficients is left for future investigation.

We still have the parameter *ζ* to discuss. The evolution equation of *S* can be written as

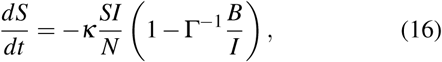

Where

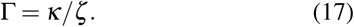

The ratio *B/I* indicates how many individuals in the cluster are typically interacting with one infected. Indeed, if we set *ζ* to zero, there is no bound on the blob-size, and a blob of one-million individuals in contact with a single infected case would not make sense physically. This is precisely the flaw in the SIR model (the well-mixedness hypothesis) that we want to improve upon in the SBIR model. The combination of the *κ* and *ζ* term will allow the system to relax to an equilibrium value Γ of the typical infection density per cluster. In the present investigation we will use Γ not only to set the ratio of *κ* and *ζ*, but also to set the initial condition for the ratio *B/I*.

We have thus given physical interpretations of the model parameters *κ, ζ*. In the following we will show how the SBIR model compares to realistic data for constant coefficients.

## III. COMPARISON WITH DATA ON CONFIRMED INFECTIONS

In this section we compare the results of our model to recent data on the epidemic. The number of daily confirmed new infected cases is obtained from the Johns Hopkins University data-base^16^. This quantity is related to the source term in equation (9),

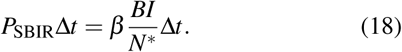

With *N*^∗^ = *B* + *I* + *R* and Δ*t* = 1 day. We have chosen to compare to several European countries where the lock-down is applied and Sweden, a country who has only applied “mild” restrictions on its population.

Our procedure is the following. We use for the parameter *γ* = 0.1^22,23^ and determine in the data the time *t* = 0 where the prevalence of daily confirmed new infected cases exceeds *P*(0) = 1·10^−6^. During the early growth phase of the epidemic, the data is fitted by an exponential, thereby determining *λ* ^+^ = *β*− *γ*. We use *I*(0) = *P*(0)*/β* and the value of *B*(0) is adjusted to match the height of the peak *P*(*τ*). The value of the cluster diffusion *κ* is varied to get a best fit on the decay phase. We note that in the phase of the epidemic we focus on, the influence of the parameter *ζ* on the results is weak compared to the influence of the choice of *κ*. It is chosen so that the initial ratio *B*(0)*/I*(0) is equal to *κ/ζ*. Finally, we impose *R*(0) = 0.

The results of the integration of the SBIR model are compared in Fig. 3 to the data, and the parameters are indicated on each plot. The general agreement is rather encouraging. It is observed that for all countries the value of *β* is of order 0.3. The value of the diffusivity varies within the subset of considered countries in the range *κ* ∈ [4.5·10^−2^, 7.5·10^−2^] and the *ζ* parameter is typically 2 to 3 orders of magnitude smaller. For all countries a revival of the epidemic starts some time after the lock-down period, which is not reproduced by the model in its present form, and the reason for this is discussed in the conclusion. The lock-down period is indicated in Fig. 3 by a shaded area. We have used an objective definition for the dates of this lock-down period using mobility data from Google^20^, given in appendix D.

**FIG. 3.**
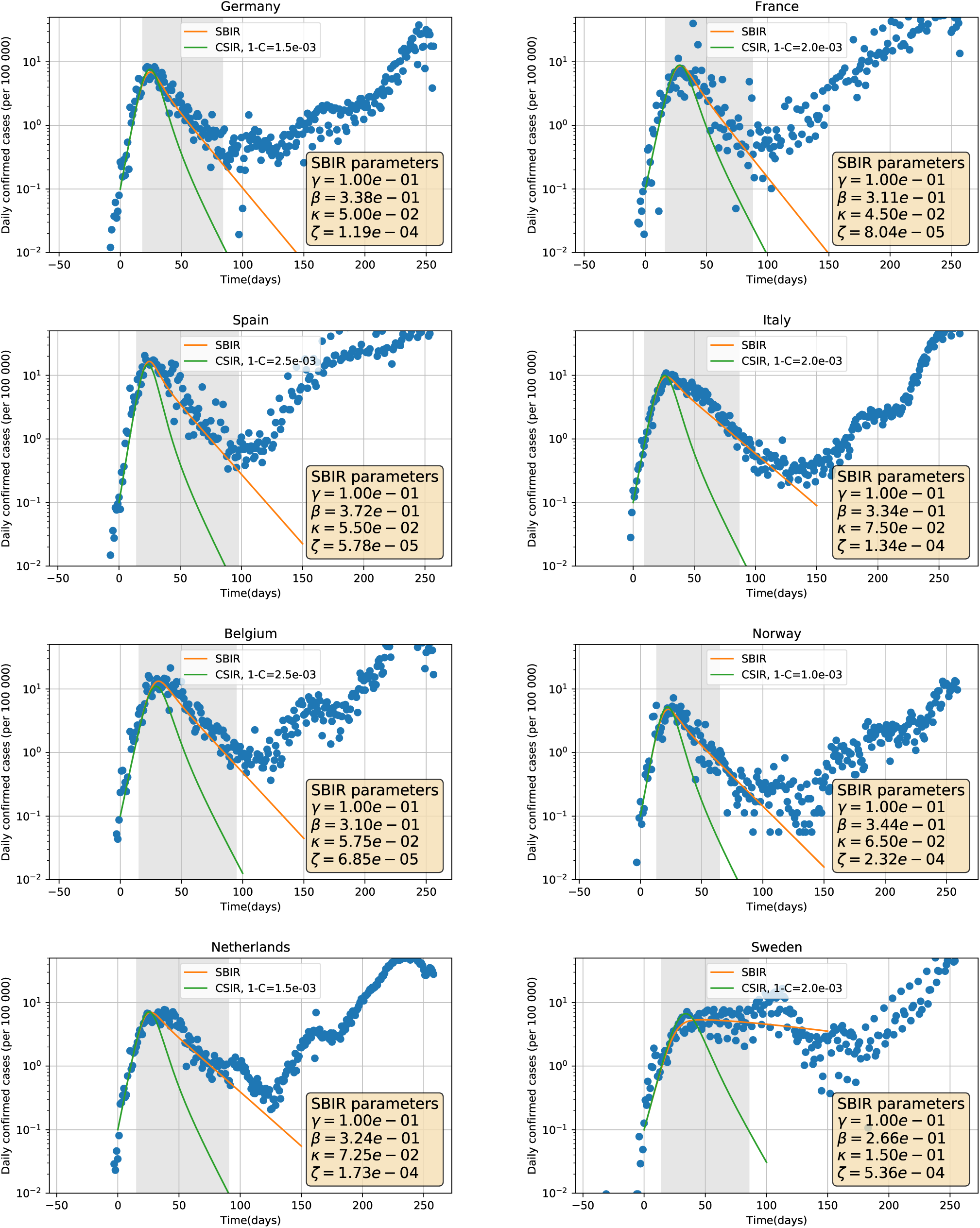
Comparison of SBIR and CSIR results, with data from JHU^16^ for 8 European countries. The data represent the reported daily new infections. The shaded region corresponds to the lock-down period in the different countries.

Also shown in the figure for comparison are the CSIR results. We have again fitted the initial growth exponent, and we have adjusted the value of the cross-immunity *C* to reproduce the peak of infection. To obtain a correct fit we need to use a value *C* ≈ 0.999, which shows that cross-immunity cannot explain the global evolution for the SARS-CoV-2 epidemic. Furthermore, the decay exponent *λ*^−^ gives fairly good agreement for *SBIR*, 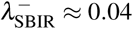(which is directly related to the value of the diffusivity parameter *κ*), whereas the CSIR decay is closer to 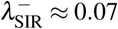, which largely underestimates the duration of the epidemic.

We also compare fhe SBIR model to a country where lock-down restrictions were very light, Sweden. It is observed that the model also describes the behaviour of the data in this country, but with a value of *κ* more than twice larger than in countries with severe lock-down restrictions. The spatial diffusion of the blob in which the ensemble of cluster evolves is in Sweden thus, according to our model, twice as large as in the other considered countries.

## IV. ESTIMATING THE REPRODUCTION NUMBER

The evolution of the number of new infected individuals can also be quantified by the reproduction number *ℛ*_*t*_. We recall that the initial value of the reproduction number within the SIR or SBIR models is defined by *ℛ*_0_ = *β/γ*.

We can rewrite the *I*-equation of the SBIR model [Eq. (11)] as

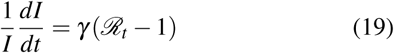

where the instantaneous reproduction number is given by the expression

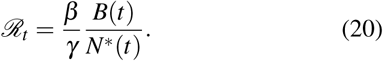

Since both *B* and *N*^∗^ evolve in time, this quantity will also vary. Initially, since *R, I≪ B*, we have *ℛ*_*t*_≈*ℛ*_0_.

In Fig. 4, we show the evolution of *ℛ*_*t*_ obtained from the SBIR-integration. To compare with data we use the local logarithmic derivative of the newly infected cases *P* to estimate *ℛ*_*t*_

**FIG. 4.**
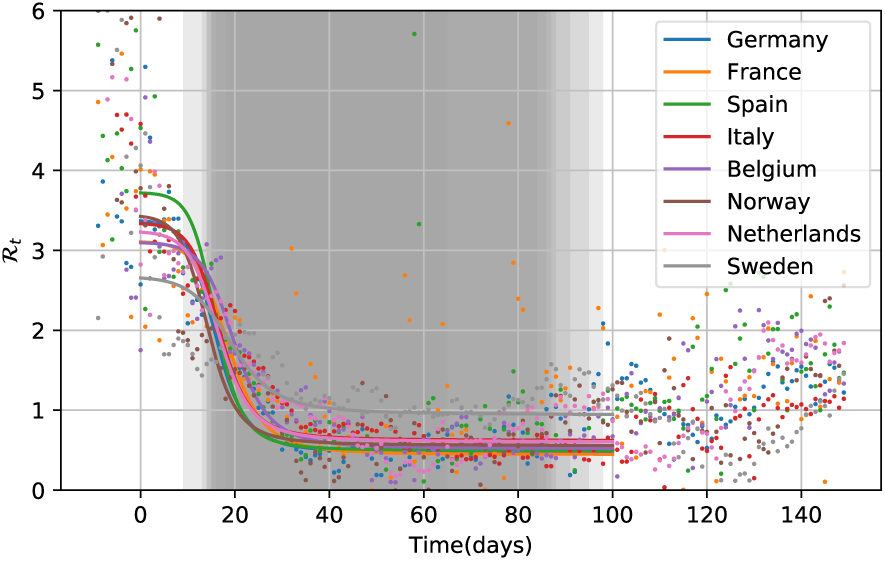
Comparison of the SBIR results for ℛ_*t*_ for 8 European countries. The shaded region is the superposition in transparency of the lock-down periods in the considered countries.

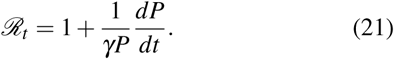

The use of *P* instead of *I* in Eq. (19) is justified as long as *B/N*^∗^ is a slowly varying quantity. In order to reduce the noise in the value of the derivative, data is preprocessed using a two-week moving average.

We see that the values of *ℛ*_*t*_ from SBIR do agree relatively well with the data. Around *t* = 0 the scatter is very large, which is in part caused by the quality of the data which depends among others on the number of tests. During lockdown, the value of *ℛ*_*t*_ drops from its initial value which varies for the considered countries around 3− 4, to a value around 0.5. The rapid decay of *ℛ*_*t*_ is present in both the data and the model estimates. The shape of the temporal evolution is close to a hyperbolic tan shape, as used in a recent modeling investigation of the COVID-19 epidemic^8^. However, there seems to be a time-delay between the two values. This might stem from the modeling approach, as no delay is introduced here between exposition and infection. The SBIR model, which is a direct improvement of the SIR model could possibly reproduce this when an E-compartment is added as in classical SEIR models.

Finally, the relation between the value of the decay exponent *λ*^−^ observed during lockdown in Fig. 3 and *ℛ* can be derived when a pure exponential is assumed,

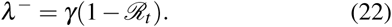

Therefore, using Eq. (11), we have for SBIR when the blob-size is determined by the diffusion of *B*,

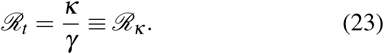

We could call *ℛ*_*κ*_ the macroscopic or global reproduction number, whereas the classical quantity *ℛ*_0_ = *β/γ* is the microscopic or local reproduction number. When *ℛ*_0_ *> ℛ*_*κ*_, the number of infected individuals in the blob will increase faster than new susceptibles can enter the blob, so that diffusion will sooner or later become the limiting factor in the spreading of the epidemic. During lock-down it is this factor which will determine the decay of the number of new infections.

## V. DISCUSSION

We have shown that the introduction of an additional compartment in the SIR model allows to take into account the presence of clusters in the evolution of an epidemic. We have shown that the SBIR model reproduces the decay-exponent of new infections during lockdown without the need to vary in time the model parameters over a duration exceeding 100 days.

The results in the present paper have to be understood as an illustration of the potential of the approach. A more comprehensive study should take into account the increase in the number of tests and its consequences on the number of new infected cases^24^. Also, we have deliberately chosen in the present work to consider the simplest model, SIR, and to add the effect of clustering upon this model. This allows to assess most easily the refinement. However, the nature of the present subdivision of compartments is so simple that most existing compartmental models, such as the SEIR model, can be modified in a similar manner.

The SBIR model shows that the close-to-universal decay exponent of the number of newly infected during lockdown, as observed in the data, can be linked to the cluster-diffusion. We also show that, during lockdown, a highly infectuous disease can be characterized by a global reproduction number *ℛ*_*κ*_ = *κ/γ*, which overrides the classical local reproduction number *ℛ*_0_ = *β/γ*.

We also show that the SBIR model in its present form reproduces observations in countries during lock-down, but no longer reproduce observations long time after restrictions are weakened. We trace back the origin of this different behavior to two reasons. The main reason is that we have assumed a constant diffusion coefficient *κ* during lock-down and we have not increased this coefficient at the end of the lock-down period. Using realistic data on mobility from mobile network providers (as used here to determine the period of lockdown) to modulate *κ*, would allow the blob to increase more rapidly and to allow the epidemic to rapidly generate new clusters.

Another reason is the fact that in the SBIR model, as formulated here, all the recovered individuals *R* are counted in the blob-population *N*^∗^. This is plausible in countries during lock-down, where clusters are fixed in space, hence will saturate and suffocate when *R* increases. In countries where the clusters not only expand, but also relocate, the epicenter of the epidemic will move spatially and leave behind regions where a large part of the population has been touched by the epidemic, but where the disease is less active. The way in which we can model these two distinct phenomenawill be discussed and assessed in a future investigation.

## Data Availability

All data are available on demand ot the authors.

## DATA AVAILABILITY STATEMENTS

The data that support the findings of this study are openly available from the Johns Hopkins University data-base^16^. Mobility data used in appendix D are openly available from^20^.

## ACKNOWLEDGMENTS

The authors thank Samuel Alizon (MIGEVEC - CNRS) and Loïc Méès (LMFA - CNRS) for stimulating discussions, as well as Benjamin Pillot (UMR Espace-Dev 228) for developing the Pycovid-19 package^25^ used to access data of the JHU-database and for useful advices. We also thank Pascal Roy (LBBE UMR 5558) and Vitaly Volpert (ICJ UMR 5208) for their helpful advices.

## Appendix A: Analytical determination of the decay exponent in SIR

The decay exponent in the SIR model can be approximated analytically. We define *t* = *τ* the time where the number of infected *I* peaks (*dI/dt* = 0). We observe exponential increase and decay before and after this time, respectively (see Fig. 2),

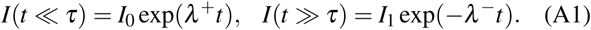

For short times *S* ≈ 1 and for very long times, *I ≪ R, S*. Therefore

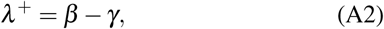

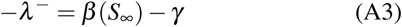

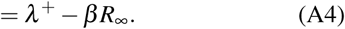

In order to determine *λ*^−^, we need therefore the total number of recovered, *R*_∞_, given by

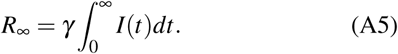

Since we assume exponential growth for short time, and exponential decay for long times, respectively, continuity at *t* = *τ* allows to write,

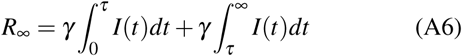

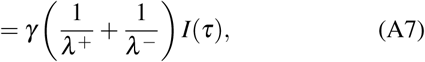

which expresses *R*_∞_ as a function of a new unknown, *I*(*τ*), the peak-value of *I*. Considering the *I*-equation,

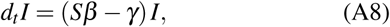

the term in brackets vanishes for *S* = *γ/β*. We know that *S* = 1 −*I* −*R*, we also have that in the initial exponential phase

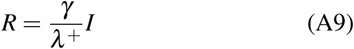

and therefore

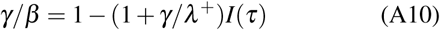

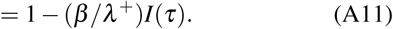

so that

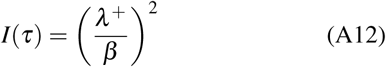

Which yields an expression for *I*(*τ*). We have the relations (A4), (A7), (A11) for the unknown quantities, *I*_*τ*_, *R*_∞_ and *λ*^−^, and solving this system yields

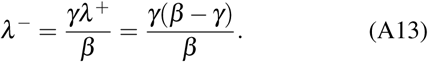

Note that for typical values, for instance *γ* = 0.2; *β* = 0.4, this yields *λ*^−^ = 0.1. Numerical integration yields *λ*^−^ = 0.119. This error of 20% is due to the assumptions of the two exponentials near the peak, where the rounding of the integrals significantly contributes to the error.

The quantity that we will consider in the comparison with data is the daily confirmed new infected *P*Δ*t*, which is associated with the source term in the *I*-equation,

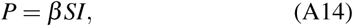

where in our system, where the time-unity is days, Δ*t* = 1.

Interesting relations, when substituting this value for *λ*^−^ in the equations are

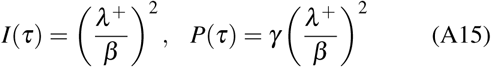

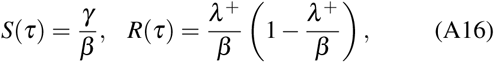

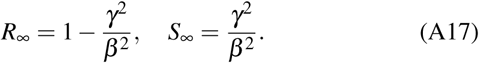

## Appendix B: Exponent in SIR with Cross immunity (CSIR)

Let us now consider the case of a partially immune population,

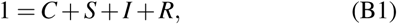

where *I*(0)*/S*(0) and *R*(0)*/S*(0) are small (as in the previous case), so that *S*(0) ≈ 1 −*C*. The *I*-equation reads

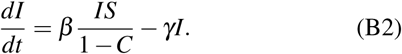

For short times, we have thus, since *S* ≈ 1 −*C*,

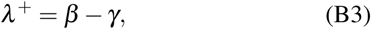

as for SIR. The growth of *I* halts at time *τ* when *dI/dt* = 0, so that we find for *S*(*τ*)

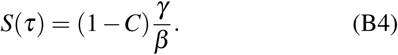

We find then for the different quantities, following the same reasoning as in the previous section,

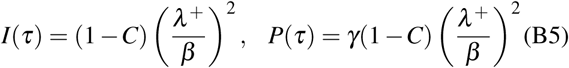

and for the exponential decay-rate,

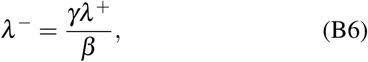

as for *SIR*.

What we learn from this is that, cross-immunity or not, if we measure *λ* ^+^, initially, *λ*^−^ is not influenced for a given *γ*. Only the height of the peak of the number of infected *I*, and of the number of daily confirmed cases *P* will be multiplied by 1 −*C*.

## Appendix C: Exponent in SBIR

Let us now consider the SBIR model. The *I* equation of SBIR reads,

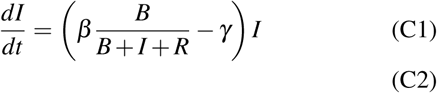

Let us consider the situation where the number of infected individuals grows faster that the number of individuals in the blob. The growth of the number of infections is then bounded by the source-term in the blob-evolution equation, and *β IB/N*\* *≈ κIS/N*. In this saturated phase, the *I*-equation becomes then,

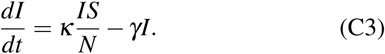

If, as during the first wave of the COVID-19 epidemic, the global infection rate is still low and *S/N* ≈ 1, it is immediately found that

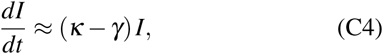

so that exponential decay is observed with a decay exponent *λ*^−^ = *γ − κ* for *κ < γ*, or a slow increase if *β < κ < γ*. The reproduction number is in this phase therefore determined by the spatial spreading, so that the value of *ℛ*_*t*_ evolves from its local, microscopic value *ℛ*_0_ = *β/γ* to its diffusion determined value *ℛ*_*κ*_ = *κ/γ*,

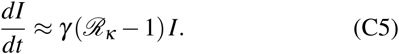

Let us evaluate when this is the case. Let us rewrite the *I*-equation as

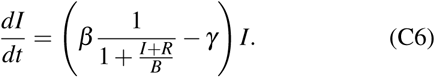

We will again assume exponentials as in Fig. 2. At short times, since *B≫ I* + *R*, we have the same increasing exponential as in SIR and CSIR, with *λ* ^+^ = *β*− *γ*. This increase will continue as long as *I* + *R≪ B*. Since *R* is in the exponential phase proportional to *I* (as for SIR), we have

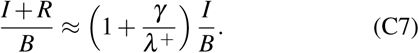

Let us therefore focus on the quantity *I/B* to see whether this quantity will be negligible for long times,

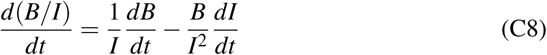

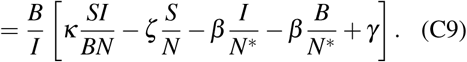

Considering the initial phase, where *I/B* ≪ 1, and estimating the order of magnitude of the different terms, it is observed that the leading order contributions of this equation are the last two terms in brackets, yielding,

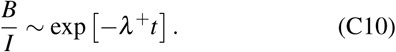

Within a finite time, the blob will thus contain a considerable number of infected and recovered individuals, so that there is no more room for additional infected.

## Appendix D: Use of Google mobility-data to determine lock-down dates

We use the mobility data provided by Google^20^ to determine the dates of the lock-down in the different countries. Fig. 5 shows the variations of mobility compared to a reference-value measured mid-january for the six countries considered in this work. The data used here corresponds to the retail and recreation data. Several other quantities are reported by Google, but most reproduce the same trend. Since we only use this data to illustrate the duration of the lockdown and do not use this to change the model-coefficients, the precise choice does not seem very important.

**FIG. 5.**
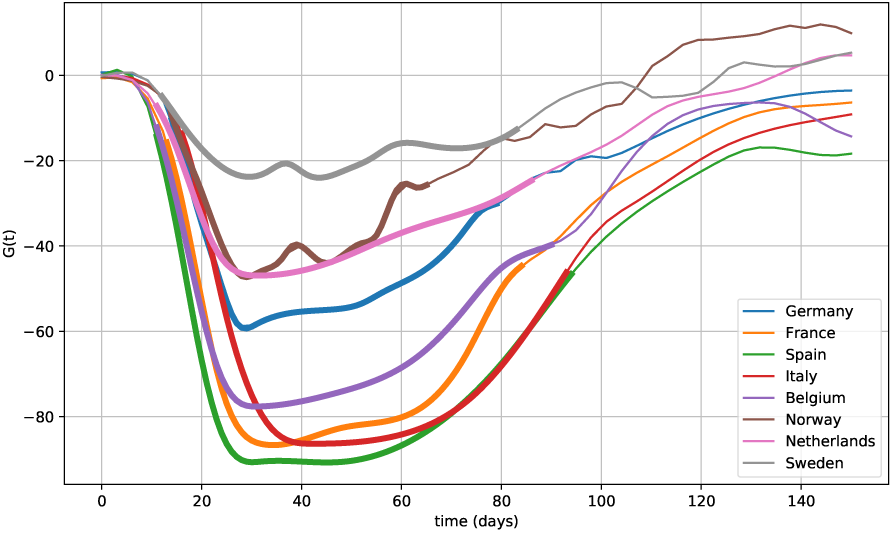
Estimation of the lock-down dates from Google mobility reports. Thick lines represent the lock-down period determined for each country.

We have smoothened the data to get rid of high-frequency variations. We call the smoothened variable **G**(*t*). The general trend shows that during the epidemic the mobility drops to a value depending on the restrictions in the different countries. An approximate plateau value is observed. We take for each country the minimum value of this plateau as the reference mobility-level during lock-down, indicated by **G**_0_. The beginning of lock-down is then determined for the time when for the first time **G**(*t*) *<* 1.15 **G**_0_ and the end, the first later date where **G**(*t*) *>* 1.5 **G**_0_, as can be seen on Fig. 5. These begin and end dates are used to indicate the lock-down periods as a shaded region in Fig. 3.

We note that this approach could allow to modulate the model coefficients. Since this will hinder the analysis of the present approach and since the assessment of the SBIR model is the main goal of the present work, this modulation is not implemented here. However, if a second wave of infections is to be reproduced, changing the model coefficients, or the dynamics of the *R*-individuals will become compulsory. Realistic mobility data can allow to get a handel on the way in which to modify the coefficients.

## References

1 William Ogilvy Kermack and Anderson G McKendrick. A contribution to the mathematical theory of epidemics. Proceedings of the royal society of london. Series A, Containing papers of a mathematical and physical character, 115(772):700–721, 1927.

2 Gemma Massonis, Julio R Banga, and Alejandro F Villaverde. Structural identifiability and observability of compartmental models of the covid-19 pandemic. arXiv preprint arXiv:2006.14295, 2020.

3 Vincenzo Capasso and Gabriella Serio. A generalization of the kermack-mckendrick deterministic epidemic model. Mathematical Biosciences, 42(1-2):43–61, 1978.

4 Nicolas Bacaër. Un modèle mathématique des débuts de l’épidémie de coronavirus en france. Mathematical Modelling of Natural Phenomena, 15:29, 2020.

5 Stéphane Derrode, Romain Gauchon, Nicolas Ponthus, Christophe Rigotti, Catherine Pothier, Vitaly Volpert, Stéphane Loisel, Jean-Pierre Bertoglio, and Pascal Roy. Piecewise estimation of R0 by a simple SEIR model. Application to COVID-19 in French regions and departments until June 30, 2020. Working paper or preprint f 02910202, August 2020.

6 Herbert W. Hethcote. The mathematics of infectious diseases. SIAM review, 42(4):599–653, 2000.

7 Fred Brauer, Carlos Castillo-Chavez, and Carlos Castillo-Chavez. Mathematical models in population biology and epidemiology, volume 2. Springer, 2012.

8 Kevin Linka, Mathias Peirlinck, and Ellen Kuhl. The reproduction number of covid-19 and its correlation with public health interventions. Comput. Mech., 66:1035, 2020.

9 Nicole Mideo, Samuel Alizon, and Troy Day. Linking within-and betweenhost dynamics in the evolutionary epidemiology of infectious diseases. Trends in ecology & evolution, 23(9):511–517, 2008.

10 James Dickson Murray, E Ann Stanley, and David L Brown. On the spatial spread of rabies among foxes. Proceedings of the Royal society of London. Series B. Biological sciences, 229(1255):111–150, 1986.

11 Marcelo Kuperman and Guillermo Abramson. Small world effect in an epidemiological model. Physical Review Letters, 86(13):2909, 2001.

12 Maia Martcheva. An introduction to mathematical epidemiology, volume 61. Springer, 2015.

13 Hazhir Rahmandad and John Sterman. Heterogeneity and network structure in the dynamics of diffusion: Comparing agent-based and differential equation models. Management Science, 54(5):998–1014, 2008.

14 David W Berger, Kyle F Herkenhoff, and Simon Mongey. An seir infectious disease model with testing and conditional quarantine. Technical report, National Bureau of Economic Research, 2020.

15 Qingchu Wu, Xinchu Fu, Michael Small, and Xin-Jian Xu. The impact of awareness on epidemic spreading in networks. Chaos: an interdisciplinary journal of nonlinear science, 22(1):013101, 2012.

16 Ensheng Dong, Hongru Du, and Lauren Gardner. An interactive web-based dashboard to track covid-19 in real time. The Lancet infectious diseases, 20(5):533–534, 2020.

17 Odo Diekmann, Johan Andre Peter Heesterbeek, and Johan AJ Metz. The legacy of kermack and mckendrick. Publications of the Newton Institute, 5:95–115, 1995.

18 Tiberiu Harko, Francisco SN Lobo, and MK Mak. Exact analytical solutions of the susceptible-infected-recovered (sir) epidemic model and of the sir model with equal death and birth rates. Applied Mathematics and Computation, 236:184–194, 2014.

19 Pieter Trapman, Frank Ball, Jean-Stéphane Dhersin, Viet Chi Tran, Jacco Wallinga, and Tom Britton. Inferring R 0 in emerging epidemics—the effect of common population structure is small. Journal of The Royal Society Interface, 13(121):20160288, August 2016.

20 COVID19 mobility reports Google. https://www.google.com/covid19/mobility. Accessed: 2020-07-03.

21 COVID19 mobility trends Apple Maps. https://www.apple.com/covid19/mobility. Accessed: 2020-07-03.

22 Huaiyu Tian, Yonghong Liu, Yidan Li, Chieh-Hsi Wu, Bin Chen, Moritz UG Kraemer, Bingying Li, Jun Cai, Bo Xu, Qiqi Yang, et al. An investigation of transmission control measures during the first 50 days of the covid-19 epidemic in china. Science, 368(6491):638–642, 2020.

23 Yinon M Bar-On, Ron Sender, Avi I Flamholz, Rob Phillips, and Ron Milo. A quantitative compendium of covid-19 epidemiology. arXiv preprint arXiv:2006.01283, 2020.

24 Thomas Sauter and Maria Pires Pacheco. Testing informed sir based epidemiological model for covid-19 in luxembourg. Working paper or preprint https://www.medrxiv.org/content/early/2020/07/25/2020.07.21.20159046, 2020.

25 Benjamin Pillot and Louis Gostiaux. Pycovid-19 python library. https://framagit.org/benjaminpillot/covid-19.

